# Viral loads of Delta-variant SARS-CoV2 breakthrough infections following vaccination and booster with the BNT162b2 vaccine

**DOI:** 10.1101/2021.08.29.21262798

**Authors:** Matan Levine-Tiefenbrun, Idan Yelin, Hillel Alapi, Rachel Katz, Esma Herzel, Jacob Kuint, Gabriel Chodick, Sivan Gazit, Tal Patalon, Roy Kishony

## Abstract

The BNT162b2 vaccine showed high real-life effectiveness both at preventing disease and in reducing viral loads of breakthrough infections, but coincidental with the rise of the Delta-variant SARS-CoV2, these protective effects have been decreasing, prompting a third, booster, vaccine inoculation. Here, analyzing viral loads of over 11,000 infections during the current wave in Israel, we find that even though this wave is dominated by the Delta-variant, breakthrough infections in recently vaccinated patients, still within 2 months post their second vaccine inoculation, do have lower viral loads compared to unvaccinated patients, with the extent of viral load reduction similar to pre-Delta breakthrough observations. Yet, this infectiousness protection starts diminishing for patients two months post vaccination and ultimately vanishes for patients 6 months or longer post vaccination. Encouragingly, we find that this diminishing vaccine effectiveness on breakthrough infection viral loads is restored following the booster vaccine. These results suggest that the vaccine is initially effective in reducing infectiousness of breakthrough infections even with the Delta variant, and that while this protectiveness effect declines with time it can be restored, at least temporarily, with a booster vaccine.

---

The Pfizer/BioNTech BNT162b2 vaccine has demonstrated high efficacy^1,2^ in preventing disease as well as real-life effectiveness for prevention of infection, disease, hospitalization, severe disease and death^3–5^. Furthermore, the vaccine has also been shown to reduce viral load for breakthrough infections (BTI), suggesting not only lower severity of disease, but also reduced infectiousness^6–8^. Indeed, national vaccination campaigns were followed by reduced infection rates, with the effect extending beyond the individual vaccinees to the community level^9–13^, giving rise to hopes for herd immunity and disease eradication. In Israel, a rapid national vaccination campaign (over 50% inoculated within 3 months) was followed by near-complete eradication of new COVID cases^14^.

Yet, despite the wide vaccination campaign and its initial dramatic impact, since mid-June, a new surge has been observed in several highly-vaccinated countries, including Israel. This increased infection rate, and especially the increased rate of BTI, can be attributed to two separate factors: waning of the vaccine induced immune response, and/or inherently lower immune response against the Delta variant (B.1.617.2)^15–17^, which became the dominant variant during the current surge. Considering the time since vaccination, studies have shown waning efficacy in protection against infection after 6 months^2,16,18,19^. Nevertheless, only a slight decline was observed in preventing severe disease and its consequences^20,21^. Considering differential effectiveness against the Delta variant, reports vary substantially as to whether, and to what extent, the vaccine is effective in reducing viral loads of BTI with the Delta variant^8,22–32^.

Given this uncertainty, in order to restrain the current wave of the pandemic, Israel, followed by the US and the UK, has decided to offer and recommend a third dose (booster) to people for whom 5 months have passed since their second shot^33^. The booster shot was first offered to people above 60 and then gradually extended to younger age groups. First signs of real-world data indicate that the booster shot is effective in preventing infections^34^, alongside results from a booster study in primates against Beta variant with the mRNA-1273 vaccine regarding viral titer^35^. Yet, it is still unclear how the booster shot affects the viral load in BTI in humans and in Delta-dominant real-world settings.

In this study, we retrospectively collected and analyzed the reverse transcription quantitative polymerase chain reaction (RT–qPCR) test measurements of three SARS-CoV-2 genes - *E, N* and *RdRp* (Allplex 2019-nCoV assay, Seegene) - from positive tests of patients of Maccabi Healthcare Services (HMS). We focus on infections of adults above the age of 20 between June 28 and August 24, when Delta was the dominant variant in Israel (over 93%)^36^. Crossing this dataset with vaccination data, we identified in total 1,910 infections of unvaccinated, 9,734 BTI of 2-dose-vaccinated and 245 BTI of booster-vaccinated (Methods: “Vaccination status”, Extended Data Table 1).

Considering all of these infections (n = 11,889), we built a multivariable linear regression model for the Ct value of each of the three genes, accounting for vaccination, at different time bins prior to the infection, and for receiving the booster as well as adjusting for sex, age and calendric date. Focusing on the *RdRp* gene, regression coefficients for vaccinated over unvaccinated started with 4.1 [95% CI: 1.6-6.6] for BTI 7-30 days post 2nd vaccine dose, yet decayed over time down to 0.7 [CI: 0.1-1.3] after about 2 months (P=0.0002, Methods: change in Ct over time) and vanished to insignificant values for infections 6 months or longer post vaccination. This decline, however, was overturned following the booster shot, which was associated with an increase of 2.2 [CI: 1.6-2.9] in the Ct, corresponding to more than 4-fold reduction in viral load (Methods: “Linear regression”, Figure 1a). Similar trends were also observed for the 2 other genes, *N* and *E* (Extended Data Fig 1). While the model adjusts for sex and age, noting that due to national rollout guidelines the average age of the booster group was higher than that of the 2-shots-vaccinated group (68.2 vs 43.8 year old), we also repeated the multivariable linear regression analysis restricted to patients above the age of 50, which yielded similar results (Extended Data Fig. 1). Finally, we note that because during the current surge only a small fraction of the patients are within their initial 2 months post-vaccination period, considering the entire population as a whole, there is only a very little viral load difference between the vaccinated and unvaccinated groups (0.26 [CI: 0.02-0.51], Figure 1b).

**Figure 1.**
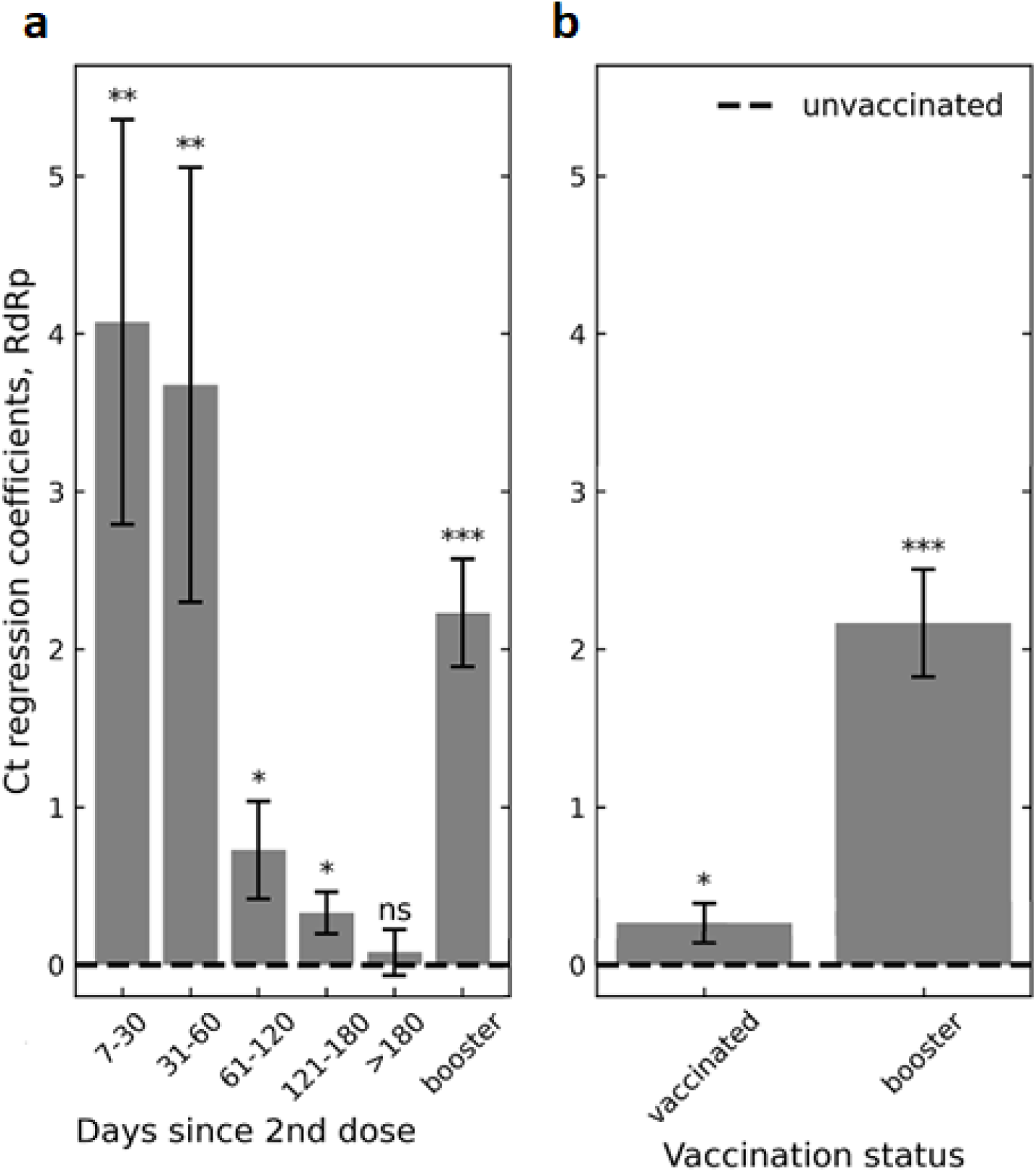
Association of infection Ct with 2-dose vaccination and with the booster. **a**, Ct regression coefficients, indicating an infection Ct relative to unvaccinated control group (dashed line), show an initial increase in Ct in the first two months following the second vaccination dose, which then gradually diminishes ultimately vanishing for infections occurring 6 months or longer post vaccination. Increased Ct is restored following the booster (right bar). Coefficients obtained by multivariate linear regression analysis adjusting for age and sex (Methods). **b**, Same model as in (a) but without binning post-vaccination times. Since most of the vaccinated population during the current surge are more than 2 months post their second vaccine shot (Extended Data Table 1), at the whole population level the average effect of the vaccine on Ct is negligible. Error bars represent one standard error. * - P-value <0.05, ** - P-value <0.01, *** - P-value <0.001. Data shown for Ct of the *RdRp* gene; for genes *N* and *E* see Extended Data Fig.1a,b.

Our results show that the vaccine is initially effective in reducing viral loads of Delta breakthrough infections, with a magnitude of 15-fold [CI: 4-53] (average over the first two months post vaccination), consistent with its initial effectiveness against pre-Delta variants^6,7^. However, this viral-load effectiveness declines with time post vaccination, significantly decreasing already after 3 months and effectively vanishing after about six months. As the Delta variant happened to appear when a large fraction of the vaccinated population was already past the initial 2 months post-vaccination, the population-wide average effect of the vaccine on Delta is negligible, consistent with and explaining reports of no difference in Ct between vaccinated and unvaccinated infected with Delta. Consistently, we found that a booster shot is associated with a regained viral-load reducing effect even during a Delta-dominated surge, suggesting restored vaccine-associated reduction in transmissibility. In total, these results suggest that the difference observed in vaccine-associated viral load reduction between the current and prior surges is associated more with immunity waning than with an inherent incapacity of reducing viral loads of BTI with the Delta variant during the immediate post second-dose or booster vaccination periods.

Our study has a number of limitations. First, although viral load is a common proxy for infectiousness, positive PCR results do not necessarily imply a viable virus and the correlation between viral loads and infectability is sensible but not fully established. Second, recent studies suggested that a faster viral load decline can be found in vaccinated compared to unvaccinated individuals with Delta infection^26,32^, offering a potential bias of Ct for samples taken long after onset of symptoms. We minimized this effect by considering only the first positive test for each patient, which is typically performed within days following symptoms^37^.

It is yet to be seen for how long the booster’s renewed effect of reducing BTI viral loads will last and whether there will be a need for additional booster shots in the future against the same variant or others to come. Nonetheless, at least in the short term, the lower viral load among booster-vaccinated Delta-BTI points to lower infectiousness which, together with other means such as social distancing and masks, could help impede the spread of the pandemic.

## Data Availability

According to the Israel Ministry of Health regulations, individual-level data
cannot be shared openly. Specific requests for remote access to de-identified community-level
data should be directed to KSM, Maccabi Healthcare Services Research and Innovation Center.

## Acknowledgments

This work was supported by the Israel Science Foundation (grant no. 3633/19 to R. Kishony and G.C.) as part of the KillCorona-Curbing Coronavirus Research Program.

## Methods

### Data collection

Anonymized SARS-CoV-2 RT–qPCR Ct values were retrieved for all of the positive samples taken between June 28 and August 24, and tested at the MHS central laboratory. For patients with multiple positive tests, only the first positive test was included. Vaccination dates for these patients were retrieved from the centralized database of MHS. Patients were excluded if their first positive sample was between the first shot and one week after the second shot. Patients with a first positive sample within the first one week following the booster shot were also excluded. Patients with positive test results before the study period were also excluded. For each test, Ct values for *E* gene, *RdRp* gene, *N* gene and the internal control were determined using Seegene proprietary software for the Allplex 2019-nCoV assay after the standard oro-nasopharyngeal swab specimen collection procedure. The same PCR model was used for all of the tests (Bio-Rad CFX96 Real-Time PCR Detection System).

### Vaccination status

Patients tested positive 7 days or more after the second shot were regarded as ‘vaccinated’. Patients tested positive 7 days or more after the third shot were regarded as ‘booster-vaccinated’. Patients tested positive less than 7 days after the second shot were excluded, as well as patients tested positive less than 7 days after their booster shot.

### Linear regression

For each of the three viral genes, we calculated the linear regression of Ct values as a function of time since second shot (length-5 onehot vector indicating 0/1 for time bins of 7<30 days, 31-60 days, 61-120 days, 121-180 days, >180 days after the second shot; all 0’s for unvaccinated), booster status (0/1), sex (0/1, female/male), age (bins of 10 years) and calendric date (number of days since June 28, 2021). Models were implemented using Python’s statsmodels library, version 0.9.0.

### Change in Ct over time

To calculate the change in Ct between post-vaccination time bins, we applied the same linear regression model as above, except replacing the length-5 onehot vector of post-vaccination time with a 3-length time-accumulated binary vector indicating [0,0,0], [1,0,0], [1,1,0], [1,1,1] for infections in unvaccinated, or 7-60 days, 61-180 days and over 180 days post the second vaccination. The coefficient of the second binary variable, indicating the *difference* in Ct between infections in the first two months and infections 2-6 months post vaccination is −3.5 [CI: (−5.4)-(−1.7)], P=0.0002 for the *RdRp* gene, −3.2 [CI: (−5.0)-(−1.4)] P=0.001 for the *E* gene and −3.3 [CI: (−5.2) - (−1.4)] P=0.001 for the *N* gene.

### Data availability

According to the Israel Ministry of Health regulations, individual-level data cannot be shared openly. Specific requests for remote access to de-identified community-level data should be directed to KSM, Maccabi Healthcare Services Research and Innovation Center.

### Ethics committee approval

This study was approved by the MHS (Maccabi Healthcare Services) Institutional Review Board (IRB). Due to the retrospective design of the study, informed consent was waived by the IRB, and all identifying details of the participants were removed before computational analysis.

## Figures

**Extended Data Figure 1.**
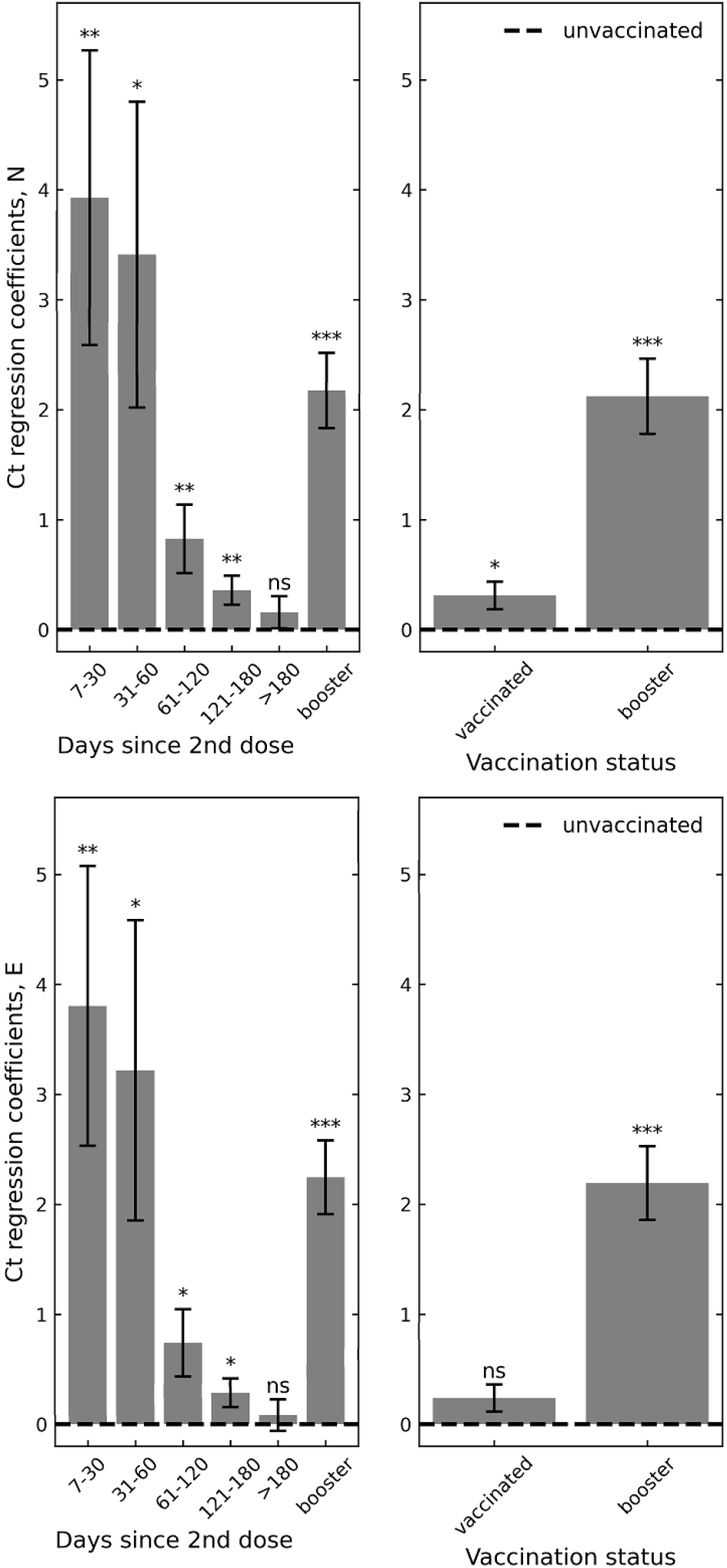
Association of infection Ct with 2-dose vaccination and with the booster, for the *N* and *E* genes. **Left**, Ct regression coefficients of the *N* gene (top) and the *RdRp* gene (bottom), indicating an infection Ct relative to unvaccinated control group (dashed line). Coefficients obtained by multivariate linear regression analysis adjusting for age and sex (Methods). **Right**, Same model but without binning post-vaccination times. Error bars represent one standard error. * - P-value < 0.05, ** - P-value < 0.01, *** - P-value < 0.001.

**Extended Data Figure 2.**
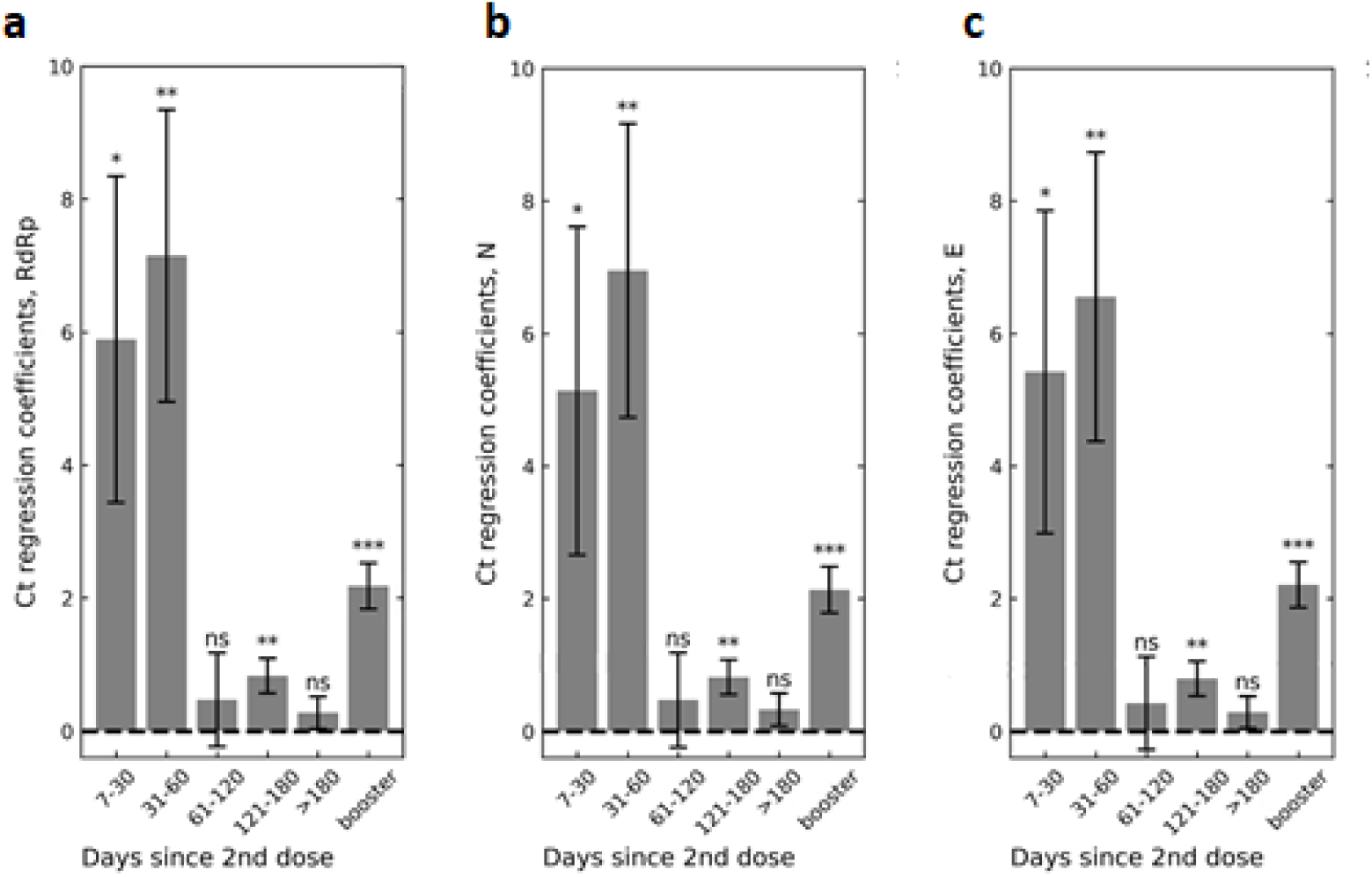
Regression coefficients for the association of Ct with 2-dose vaccination and with the booster for patients older than 50. Same model as in Figure 1 and Extended Data Figure 1, but restricting for patients 50 years or older. Ct regression coefficients are shown for the *RdRp* (a), *N* (b) and *E* (c) genes. Error bars represent one standard error. * - P-value < 0.05, ** - P-value < 0.01, *** - P-value < 0.001.

**Extended Data Table 1:** Number of infections by time since vaccination and booster shot.

| **Time since**<br>**2nd shot (days)** |  |  |  |  |  |  |
| --- | --- | --- | --- | --- | --- | --- |
| **Vaccination**<br>**status** | **7-30** | **31-60** | **61-120** | **121-180** | **>180** | **total** |
| **unvaccinated** | - | - | - | - | - | 1,910 |
| **2-dose-vaccinated** | 15 | 13 | 304 | 5,880 | 3,522 | 9,734 |
| **booster-vaccinated** | 0 | 0 | 1 | 8 | 236 | 245 |

